# Comparison of Pallidal Deep Brain Stimulation and Bilateral Pallidotomy for Medically Refractory Status Dystonicus: A Systematic Review and Meta-Analysis

**DOI:** 10.1101/2025.09.09.25335446

**Authors:** James Kelbert, Diego Soto Rubio, Jacob A. Saunders, Michael C. Kruer, Robert W. Bina

## Abstract

**Purpose:** Status dystonicus (SD) is a medical emergency that results from a primary or secondary dystonia condition and is life-threatening if untreated. Surgical intervention is indicated for swift resolution of medically refractory SD. This study looks to compare bilateral pallidotomy against deep brain stimulation (DBS) for SD treatment.

**Methods:** A meta-analysis was conducted according to PRISMA guidelines. Terms “pallidotomy”, “deep brain stimulation” and “status dystonicus” were used on January 2nd, 2024 to search PubMed, EMBASE, Scopus and Web of Science. All article data was uploaded to the systematic review software Rayyan where duplicates were removed with three independent reviewers screening remaining articles. Data was manually collected after quality assessment and analyzed using meta-analysis of proportions of pooled complications and continuous meta-analysis of one of two movement disorder scores.

**Results:** Forty-three total patients (32 DBS and 11 pallidotomy; 27 male and 16 female) were included. Average age of SD onset was 12.49 ± 8.55 years (11.95 ± 9.69 for DBS and 13.71 ± 5.30 for pallidotomy) Change in movement disorder score was 48 ± 34% overall (54 ± 33% for DBS and 34 ± 33% for pallidotomy). A subgroup meta-analysis showed a standardized mean difference of changes in movement disorder score of - 1.55 [−2.97, −0.13] for DBS and −1.35 [−2.40, −0.30] (p=0.83). A meta-analysis of proportions showed a 40% complication rate for DBS and 16% complication rate for pallidotomy (p=0.14).

**Conclusion:** There is insufficient evidence to determine if there is a difference between pallidotomy or DBS for SD treatment, but these results suggest that pallidotomy may be a valid alternative in resource-limited settings or when DBS is contraindicated, although further comparative studies are needed.

## Introduction

Status dystonicus (SD), or dystonic storm, is a rare, yet serious medical condition that affects patients of all ages with a mortality rate of 10-12.5% with children and adolescents at higher risk[1, 2]. While epidemiology is difficult to characterize, one cross-sectional study found that the annual incidence rate of SD in children was 0.05 out of every 1000 new admissions[3]. No specific diagnostic criteria exist, but SD is an emergency characterized by increasingly severe and frequent generalized or focal dystonia requiring hospitalization for treatment and management[2–4].

First-line treatment for SD is medical management with anti-spasmodic agents such as vesicular monoamine transporter inhibitor tetrabenazine and antimuscarinic benztropine[5]; however, given the severity of SD, pharmacological management is often insufficient to control symptom progression[5, 6]. In one study, only 10% of cases were resolved by pharmaceutical intervention alone[1]. Thus, neurosurgical interventions are often utilized to treat medically refractory SD. Deep brain stimulation (DBS) with targets of GPi and STN has been used to treat SD with reported success rates of greater than 92 percent[1, 7–10].

Several barriers exist that make widespread DBS implementation difficult. The implantation of a device places a financial burden on the patient, which is estimated to be $162,489 US dollars over five years[11]. This figure does not account for the additional lifetime cost for management of the implanted hardware and does not account for costs related to device malfunction or failure[11, 12]. Moreover, device access is limited worldwide for a variety of reasons including cost and long-term device management[13]. Given some of these constraints, additional treatment options are needed to address the needs of patients with SD when DBS may be contraindicated or unavailable.

Pallidotomy was among the first surgical treatments for SD[3, 12, 14]. The specific target for pallidotomy has evolved over time, with the most recent consensus target being the GPi[12, 15]. In several studies, pallidotomy was shown to be effective in treating and resolving SD, with improvement of standardized movement disorder scales indicating SD resolution in as high as 83% of patients in one study[12, 15].

Given the modulative nature of DBS along with its favorable side effect profile, it is the more preferred neurosurgical treatment for medically refractory SD, yet there is a paucity of evidence directly comparing DBS to pallidotomy in the management of SD. Most published studies are case reports and case series; given this, the present systematic review aims to compare the efficacy of the two therapies in the treatment of this condition.

## Methods

This study was a systematic review and meta-analysis following the Preferred Reporting Items for Systematic Reviews and Meta-Analyses (PRISMA) guidelines.[16] The population of interest was medically refractory SD patients who received either bilateral DBS or radiofrequency pallidotomy.

### Search Strategy and Inclusion

Database search was conducted on January 2^nd^, 2024, on PubMed, Scopus, EMBASE and Web of Science. The terms “pallidotomy”, “deep brain stimulation” and “status dystonicus” were used to search these databases using the Boolean operator AND to maximize sensitivity. A full protocol has been published under PROSPERO registration number CRD42024500543 on January 21^st^, 2024 before data extraction and analysis was conducted. The PROSPERO was updated on November 17^th^, 2024 to include further direct analysis including individual patient data.

No additional search parameters, filters, or limits were applied. Once the search was complete, all article data was uploaded to the systematic review software Rayyan[17] for the screening and selection process. Any duplicates of records and records marked as ineligible by automation tools were removed prior to the screening process. Titles and abstracts of remaining records were then screened by three independent reviewers (JK, DSR, and JS). Conflicting assessments of records were resolved by consensus.

### Inclusion and Exclusion Criteria

Study Inclusion criteria were as follows: any case report that investigated the use of DBS or pallidotomy for treatment of medically refractory SD; studies in the English language; and patients of any age diagnosed with SD of any etiology. Study Exclusion criteria were as follows: studies that include either deep brain stimulation outside of globus pallidus or lesion of other brain area (rhizotomy, thalamotomy, etc.) for treatment; studies in a language other than English; studies that do not report an objective movement disorder scale such as Burke-Fahn-Marsden Dystonia Rating Scale (BFMS), Barry-Albright Dystonia Scale (BAD), Unified Dystonia Rating Scale (UDRS), amongst any others; and abstracts, letters to the editor, editorials, meta-analysis, review studies or any other study that does not report any primary data.

### Data Extraction

After all included studies were finalized, data was manually collected from each study by searching the full text version where authors JK, DSR and JS all worked on the inclusion and exclusion of articles separately and then convened to include or exclude articles by unanimous consensus. The following data was extracted: study design, study period, number of participants, mean age, gender breakdown, age of dystonia onset, age of SD onset, SD etiology, follow-up time after surgery, surgical operation performed, associated comorbidities, pre-operative clinical outcomes (preoperative standardized movement disorder score) and post-operative outcomes (postoperative standardized movement disorder score and number of complications). For DBS patients, it was specifically noted if the SD episode was secondary to DBS failure and/or removal.

### Quality assessment

For retrospective case series and cohort studies, the Joanna Briggs Institute[18] (JBI) critical appraisal tools were utilized for the respective studies by authors JK and DSR. It was decided for ultimate inclusion for a study to score at least eight out of ten using the JBI case series checklist.

### Statistical Analysis

Statistical analysis was performed using R version 4.3.3 (R Foundation for Statistical Computing, Vienna, Austria). Change in movement disorder scale score, mean age of dystonia onset, SD onset and follow-up between groups were compared using a non-parametric Mann-Whitney-U test. Analysis of standardized mean differences in various dystonia rating scale (BFMRS, BAD, UHDRS, etc.) scores were conducted using a random effects meta-analysis where the control group utilized the pre-operative scores, and the experimental group involved the post-operative clinical outcome scores.

A random effects model was used, and heterogeneity was determined with a restricted maximum-likelihood estimator for tau-squared, Q-Profile method for confidence interval of tau-squared and tau along with an I-squared analysis. A meta-analysis of proportions was conducted to determine any differences in total number of reported complications between groups. Complications were counted if they were directly related to the process of undergoing surgery or to the surgical intervention (DBS or pallidotomy) itself. This was conducted using an inverse method with a logit transformation and Clopper-Pearson confidence intervals with the same methods of heterogeneity quantification previously mentioned.

To account for small study effects, funnel plots were generated, but imprecise quantification of publication in funnel plots of meta-analysis of proportions was acknowledged[19]. Egger’s test was performed for both meta-analyses, although it was further acknowledged that it is likely to overestimate bias within this sample when there are fewer than ten total studies.

### Grading the Evidence

Summary of findings and overall confidence in accumulated evidence was assessed using the Grading of Recommendations Assessment, Development and Evaluation (GRADE) system.[20] GRADE rates the quality of evidence across the different following domains: directness, risk of bias, consistency, precision and publication bias. Accounting for all these domains, GRADE ranks the variable into one of four categories: *high* (further research is very unlikely to change our confidence in our effect estimate), *moderate* (further research is likely to change our confidence in our effect estimate), *low* (further research is very likely to change our effect estimate), or *very low* (our effect estimate is very uncertain).

## Results

### Inclusion of Publications

Of 458 studies which resulted from the four database searches, 300 duplicate articles were removed, and 158 publications were initially screened. Of those 158, 42 met inclusion criteria and were all retrieved for a full text screening (Figure 1). Of those 42, 13 studies were included in addition to 6 studies found through backward and forward citation analysis (Figure 1) leading to a total of eighteen studies (13 DBS and 4 pallidotomy and 1 DBS and pallidotomy) with 43 total patients (27 male and 16 female; 32 DBS and 11 pallidotomy) included in the systematic review (Table 1)[12, 15, 21–36].

**Figure 1:**
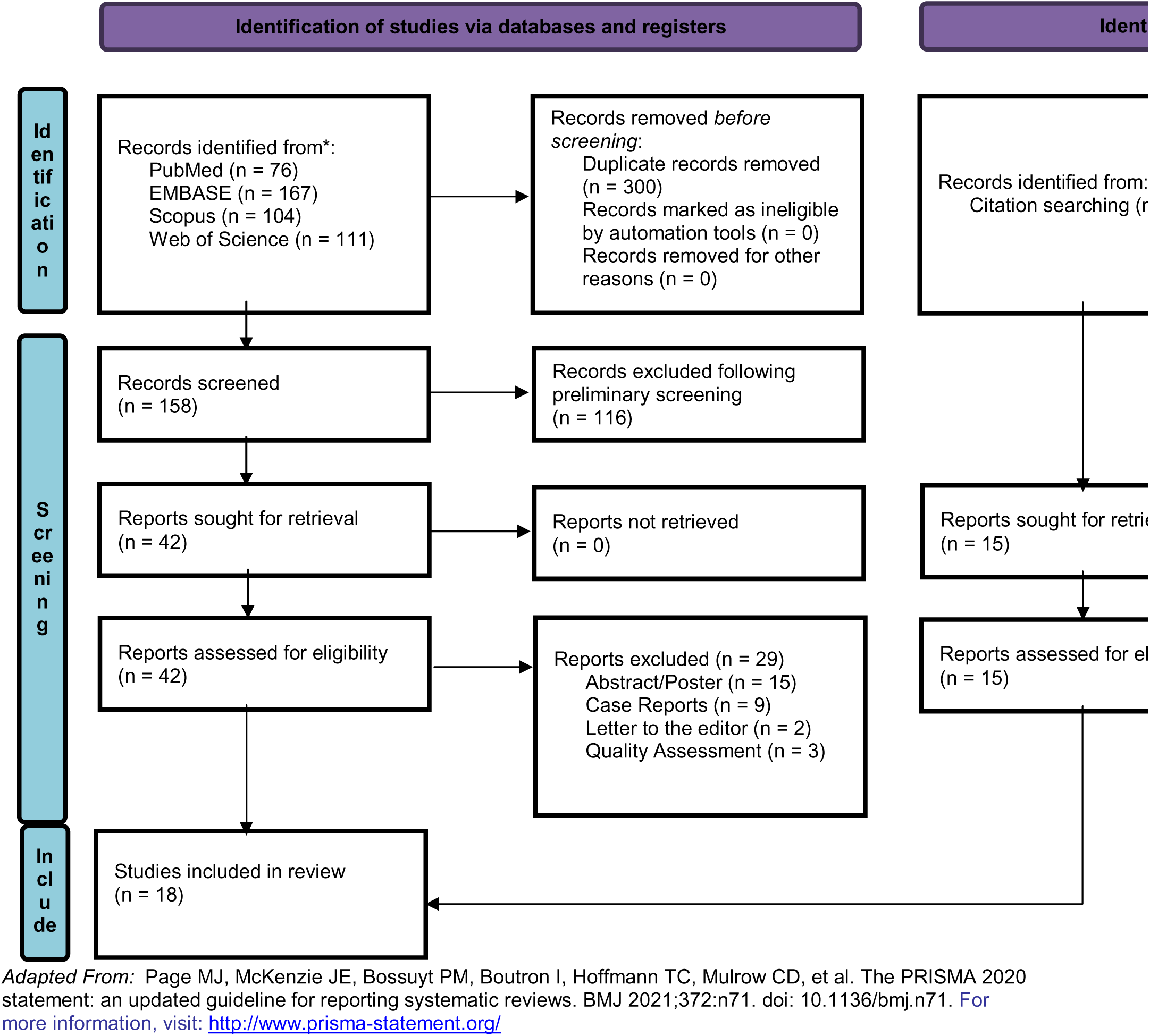
PRISMA template for all included studies.

**Table 1:** Demographic information of included case reports and series within the systematic review.

| *Authors* | *Gender* | *Age*<br>*Onset* | *Age*<br>*at SD* | *Etiology* | *Score*<br>*Utilized* | *Movement*<br>*Disorder*<br>*Score Pre-*<br>*surgery* | *Movement*<br>*Disorder Score*<br>*Post-surgery* | *Follow-up*<br>*time point*<br>*(months)* | *Modality* |
| --- | --- | --- | --- | --- | --- | --- | --- | --- | --- |
| *Mithani et al.* | F | 0.1 | 3 | SCN2A | BFMDRS<br>total | 98 | 60 | 6 | DBS |
| *Zaman et al.* | M | unknown | 7 | UBA5 | DSS | 5 | 1 | 4 | DBS |
| *Tavasoli et al.* | M | 11 | 11 | anti-NMDA receptor<br>encephalitis | BFMDRS<br>total | 98 | 42 | 9 | DBS |
| *Aydin et al.* | M | unknown | 5 | Epilepsy | BFMDRS<br>total | 106 | 20 | 15 | DBS |
| *Kovacs et al.* | M | 18 | 18 | Tardive dystonia | BFMDRS<br>total | 108 | 3.5 | 12 | DBS |
| *Honey et al.* | M | 1.5 | 10 | GNAO1 | BAD<br>total | 27 | 5 | 6 | DBS |
| *Franzini et al.* | F | 4 | 6 | Tubercular<br>meningoencephalitis | BFMDRS<br>total | 110 | 50 | 3 | DBS |
| *Borggraefe et al.* | F | 3 | 10.6 | DYT1 | BFMDRS<br>total | 75 | 5 | 14 | DBS |
| *Jech et al.* | M | 8 | 11 | Primary dystonia<br>(DYT1 negative) | BFMDRS<br>total | 41 | 3 | 15 | DBS |
| *Ben-Haim et al.* | F | 6 | 13 | DYT1 | BFMDRS<br>total | 96 | 0 | range of 12-96 | DBS |
| *Ben-Haim et al.* | M | 8 | 11 | DYT1 | BFMDRS<br>total | 28 | 4 | range of 12-96 | DBS |
| *Zorzi et al.* | M | 1 | 8 | Primary dystonia<br>(DYT1 negative) | BFMDRS<br>total | 91 | 83 | 15 | DBS |
| *Zorzi et al.* | M | 0.5 | 14 | Encephalopathy of unknown origin | BFMDRS total | 43 | 43 | 15 | DBS |
| *Zorzi et al.* | M | 3 | 10 | Primary dystonia<br>(DYT1 negative) | BFMDRS<br>total | 79.5 | 63 | 19 | DBS |
| *Lobato-Polo*<br>*et al.* | M | 41 | 41 | Idiopathic | UDRS<br>total | 56 | 0 | 43 | DBS |
| *Lobato-Polo*<br>*et al.* | M | 8 | 0.1 | Cerebral palsy | UDRS<br>total | 42 | 9 | 27 | DBS |
| *Lobato-Polo*<br>*et al.* | M | 41 | 41 | Idiopathic | UDRS<br>total | 30 | 17.5 | 15 | DBS |
| *Lobato-Polo*<br>*et al.* | M | 10 | 1 | Cerebral palsy | UDRS<br>total | 42 | 20.5 | 27 | DBS |
| *Lobato-Polo*<br>*et al.* | F | 32 | 7 | *GCH1* mutation | UDRS<br>total | 42 | 10 | 13 | DBS |
| *Levi et al.* | F | 1 | 6 | GNAO1 | BAD<br>total | 32 | 21 | 12 | DBS |
| *Levi et al.* | F | 3 | 10 | Cerebral palsy | BAD<br>total | 32 | 22 | 63 | DBS |
| *Levi et al.* | M | 6 | 14 | Cerebral palsy | BAD<br>total | 31 | 25 | 19 | DBS |
| *Levi et al.* | M | 11 | 13 | PKAN classic | BAD<br>total | 32 | 24 | 84 | DBS |
| *Levi et al.* | M | 4 | 9 | PKAN classic | BAD<br>total | 31 | 28 | 36 | DBS |
| *Levi et al.* | F | 2 | 10 | PKAN classic | BAD<br>total | 30 | 28 | 42 | DBS |
| *Levi et al.* | F | 12 | 24 | PKAN classic | BAD<br>total | 32 | 25 | 28 | DBS |
| *Zhai et al.* | F | 8 | 9 | PKAN classic | BFMDRS<br>total | 92 | 20 | 3 | DBS |
| *Elkay et al* | F | 17 | 19 | Batten's disease | BFMDRS<br>total | 100 | 62 | 7 | Pallidotomy |
| *Balas et al.* | M | 5 | 10 | HSD | BFMDRS<br>total | 116 | 47 | 48 | Pallidotomy |
| *Marras et al.* | M | 3 | 15 | Chromosomopathy | BFMDRS<br>total | 101 | 16 | 22 | Pallidotomy |
| *Marras et al.* | M | 0.1 | 19 | Epileptic<br>encephalopathy | BFMDRS<br>total | 84 | 4 | 21 | Pallidotomy |
| *Marras et al.* | M | 0.3 | 6.5 | Idiopathic bilateral<br>striatal necrosis | BFMDRS<br>total | 60 | 57 | 15 | Pallidotomy |
| *Marras et al.* | M | 0.67 | 12 | Severe neonatal<br>cerebral hypoxia | BFMDRS<br>total | 77 | 44 | 15 | Pallidotomy |
| *Levi et al.* | F | 1 | 10 | GNAO1 | BAD<br>total | 32 | 28 | 180 | Pallidotomy |
| *Levi et al.* | F | 4 | 7 | Post-infective | BAD<br>total | 32 | 31 | 12 | Pallidotomy |
| *Levi et al.* | M | 1 | 9 | Unknown | BAD<br>total | 30 | 15 | 12 | Pallidotomy |
| *Levi et al.* | M | 2 | 22 | PKAN atypical | BAD<br>total | 31 | 30 | 12 | Pallidotomy |
| *Levi et al.* | F | 1 | 19 | Cerebral palsy | BAD<br>total | 31 | 28 | 38 | Pallidotomy |
| *Levi et al.* | M | 2 | 16 | Cerebral palsy | BAD<br>total | 30 | 28 | 30 | Pallidotomy |

Abbreviations: SD: status dystonicus; SCN2A: sodium voltage-gated channel alpha subunit 2; UBA5: Ubiquitin-like modifier-activating enzyme 5; NMDA: N-methyl D-aspartate; GNAO1: G protein subunit alpha o1; DYT1: Dystonia 1 protein; GCH1: GTP cyclohydrolase 1; PKAN: Pantothenate kinase-associated neurodegeneration; HSD: Hallervorden–Spatz disease; BFMDRS: Burke-Fahn-Marsden Dystonia Rating Scale; DSS: dystonia severity scale; UDRS: United Dystonia Rating Scale; BAD: Barry-Albright Dystonia rating scale; DBS: deep brain stimulation

### Demographics

Average age of SD onset was 12.49 ± 8.55 years (11.95 ± 9.69 for DBS and 13.71 ± 5.30 for pallidotomy with a p-value of 0.15) (Table 1). Average age of dystonia onset in years was 7.57 ± 10.30 (9.72 ± 11.6 for DBS and 3.09 ± 4.63 for pallidotomy with a p-value of 0.01) (Table 1). Average follow-up was 25.86 ± 31.19 months (21.18 ± 19.35 months for DBS and 34.33 ± 47.44 months for pallidotomy with a p-value of 0.44) (Table 1). The mean change in corresponding movement disorder score was 48 ± 34% overall (54 ± 33% for DBS and 34 ± 33% for pallidotomy). While there is over a twenty-point difference, there was no statistically significant difference found in change in standardized movement disorder scale score between DBS and pallidotomy groups (Figure 2; p= 0.09).

**Figure 2:**
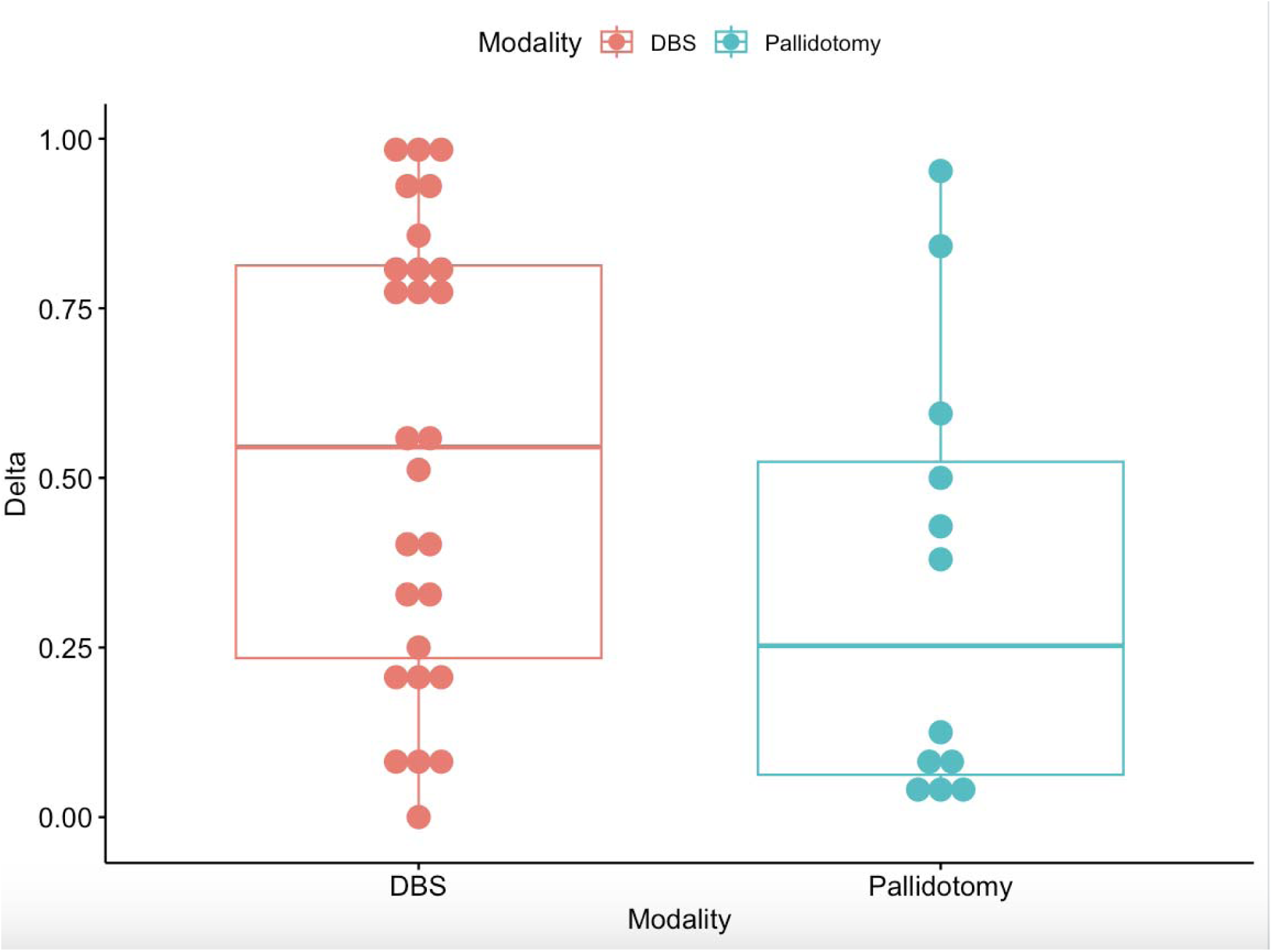
Change in normalized movement disorder scale score for DBS and pallidotomy respectively. There is no significant difference between the two groups as determined by Mann-Whitney-U test (p=0.09). Figure 2: Change in normalized movement disorder scale score for DBS and pallidotomy respectively. There is no significant difference between the two groups as determined by Mann-Whitney-U test (p=0.09).

A variety of dystonia etiologies led to SD and required surgical intervention (Table 1): six cerebral palsy (four DBS and two pallidotomy), six Dystonia 1 protein (*DYT1)*-positive dystonia (five DBS), five pantothenate kinase-associated neurodegeneration (*PKAN)* classic mutation (five DBS), four idiopathic or unknown origin (three DBS and one pallidotomy), three *DYT1*-negative dystonia (three DBS), three G protein subunit alpha o1 (*GNAO1)* mutation (two DBS and one pallidotomy), one GTP cyclohydrolase 1 (*GCH-1)* mutation (DBS), one sodium voltage-gated channel alpha subunit 2 (SCN2A) mutation (DBS), one anti-N-methyl-D-aspartate (NDMA) receptor encephalitis (DBS), one Ubiquitin-like modifier-activating enzyme 5 (UBA5), one unspecified epilepsy (DBS), one tardive dystonia (DBS), one tubercular meningoencephalitis (DBS), one Dystonia 6 protein (*DYT6)*-positive dystonia (DBS), one post-infective (pallidotomy), one unspecified chromosomopathy (pallidotomy), one *PKAN* atypical mutation (pallidotomy), one epileptic encephalopathy (pallidotomy), one idiopathic bilateral striatal necrosis (pallidotomy), one with *Hallervorden–Spatz disease* (HSD; pallidotomy), one with Batten’s disease (pallidotomy) and one severe neonatal cerebral hypoxia (pallidotomy).

### SD secondary to DBS failure

Two studies comprising four patients (two male and two female) that involved SD secondary to DBS hardware failure or removal were identified (Table 2)[30, 34]. The mean time to SD after DBS failure or removal was 6.75 ± 7.04 months. All patients had either a DYT1 or DYT6 mutation as their dystonia etiology. After repair or replacement of DBS device, all four patients were eventually extubated and had significant improvement in BFMDRS scores. Only one patient was lost to follow-up. The only reported complication was transient left hemiparesis in one patient.

**Table 2:** Status Dystonicus after Deep Brain Stimulation hardware failure and/or removal.

| *Article* | *Gender* | *Age* | *Age at SD* | *Previous DBS* | *Etiology* | *Score* | *Movement* | *Movement* | *Follow-up* |
| --- | --- | --- | --- | --- | --- | --- | --- | --- | --- |

|  | *onset* |  |  | *placement* |  | *Utilized* | *Disorder Score Pre-surgery* | *Disorder Score Post-surgery* | *(months)* |
| --- | --- | --- | --- | --- | --- | --- | --- | --- | --- |
| *Ben-Haim et al.* | F | 7 | 8 | Immediately after discharge from DBS placement | DYT1 | BFMDRS total | 49 | NA | range of 12-96 |
| *Ben-Haim et al.* | F | 7 | 11 | 1 month after bilateral lead fractures | DYT1 | BFMDRS total | 104 | 95.5 | range of 12-96 |
| *Ben-Haim et al.* | M | 8 | 11 | 2 months after DBS removal | DYT1 | BFMDRS total | 136 | 68 | range of 12-96 |
| *Oterdoom et al.* | M | 3.5 | 11 | 15 months prior | DYT6 | BFMDRS total | 119 | 79 | 36 |

### Meta-analysis Findings

Of those nineteen included studies, four (two DBS, one pallidotomy and one DBS versus pallidotomy study) were ultimately included in a meta-analysis of 25 patients (17 male and 8 female) (Table 1).

The meta-analysis generated an overall standardized mean difference in movement disorder score of −1.41 [−2.09, −0.73]. For the DBS, the score was −1.55 [−2.97, −0.13] and −1.35 [−2.40, - 0.30] for pallidotomy with a chi-square test for subgroup difference p-value of 0.83 and an I2 of 21.5% (Figure 3a). To account for small study effects, a funnel plot showed no findings indicative of publication bias with an Egger’s test of p=0.31 (Figure 4a).

**Figure 3:**
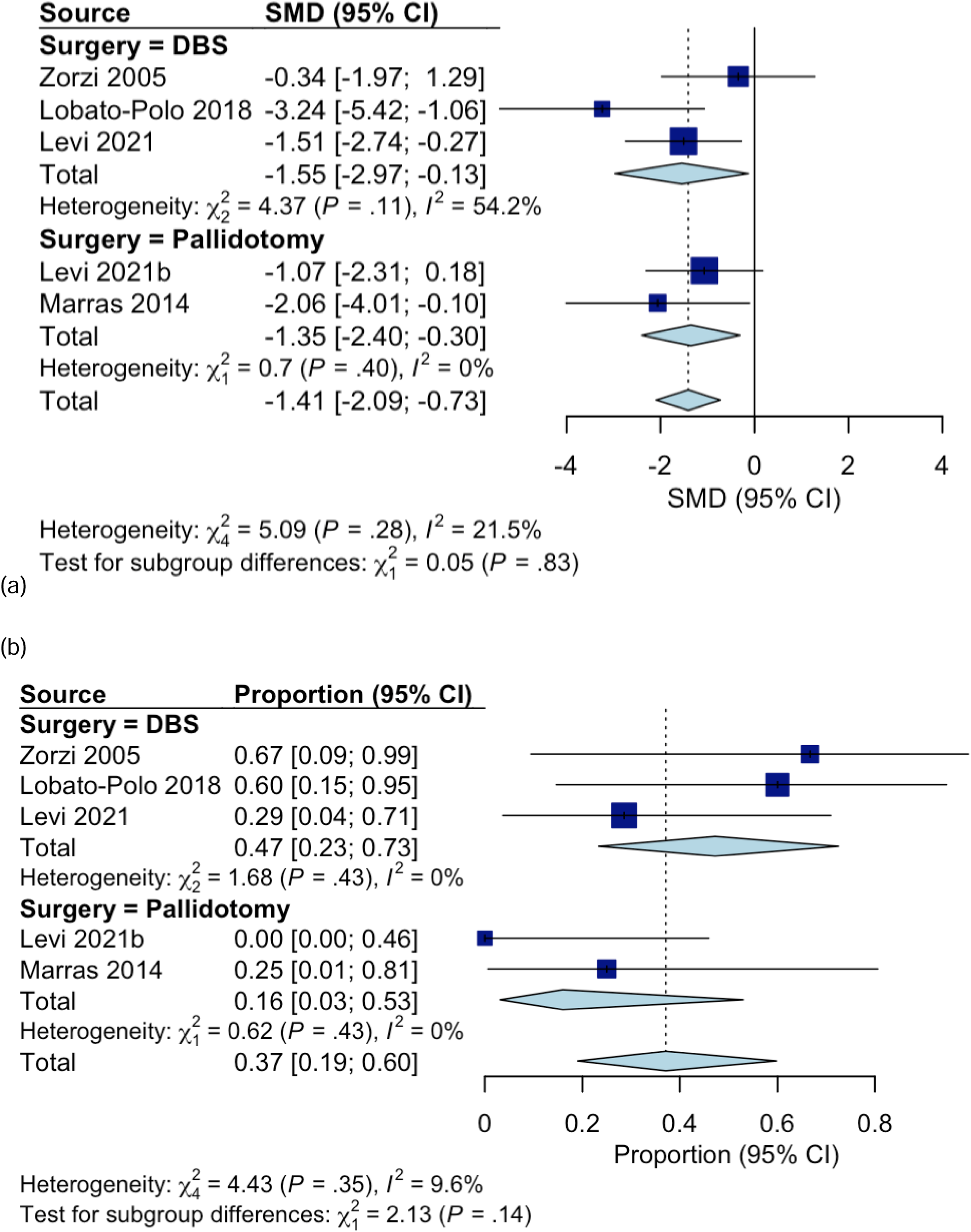
Forest plot comparing pre- and post-surgical movement disorder scores in (a) and complication rates in (b) for deep brain stimulation and bilateral pallidotomy for status dystonicus treatment.

**Figure 4:**
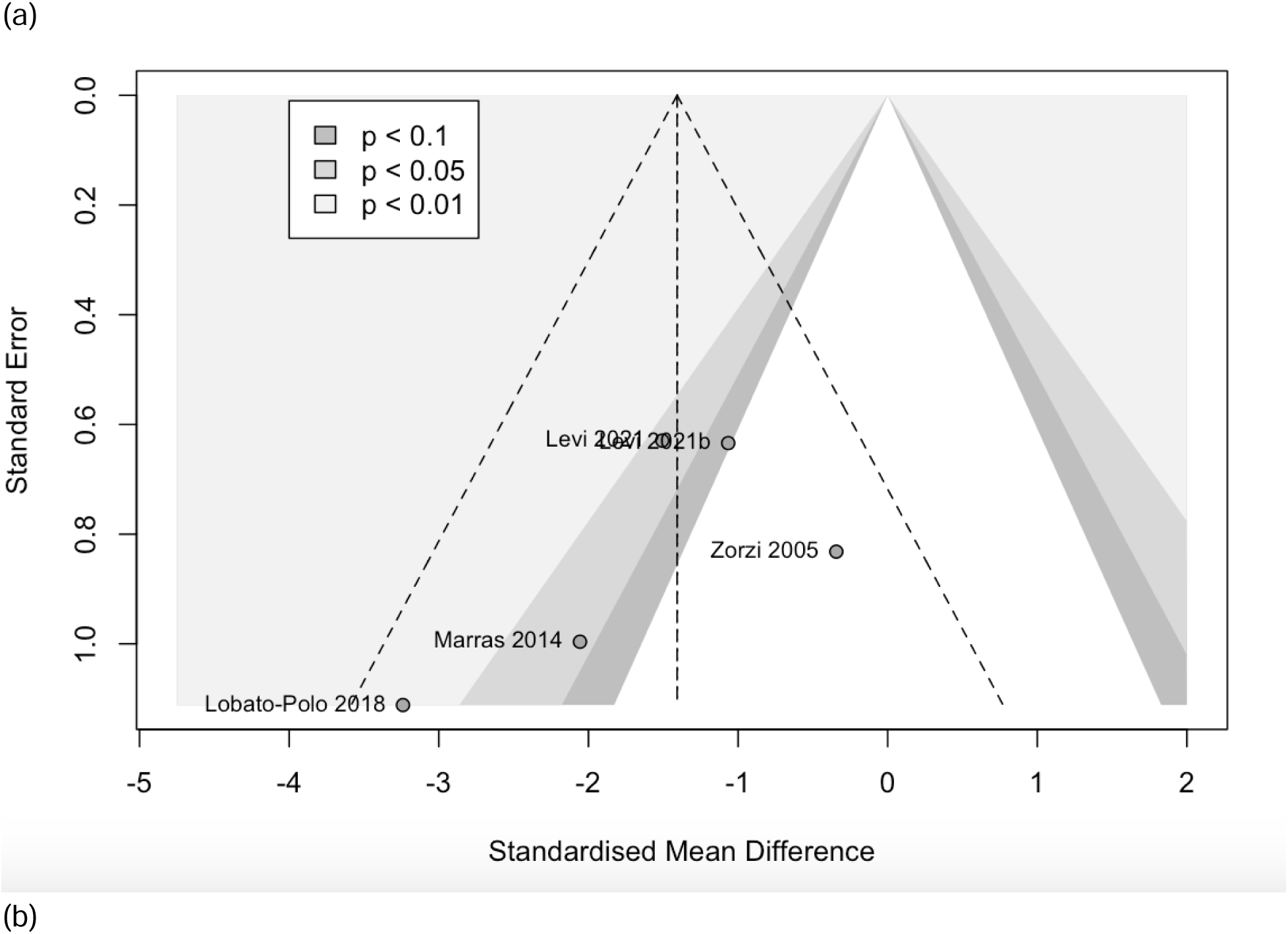
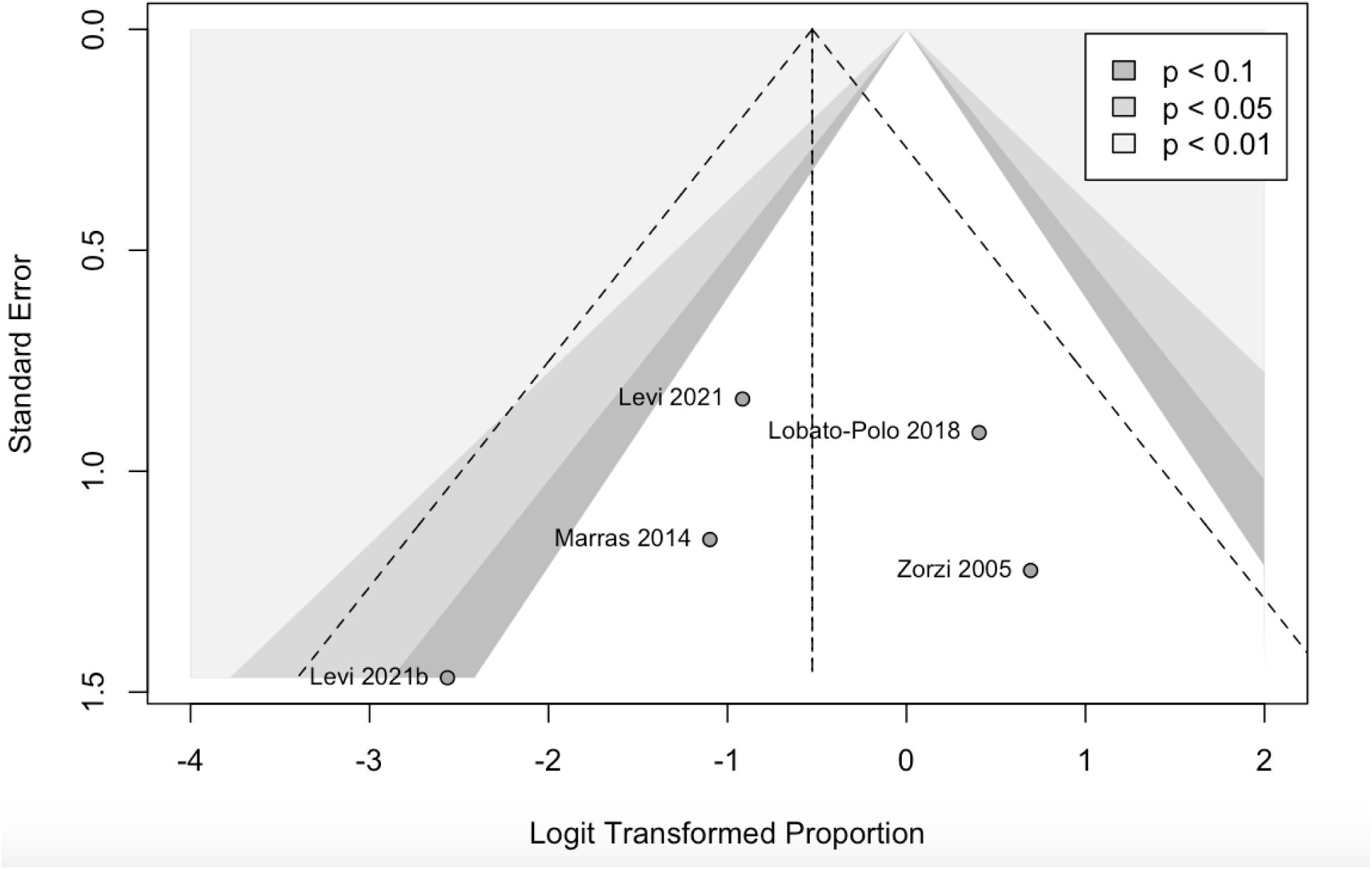
Small study effects analysis using a contour-enhanced funnel plot for standardized mean differences for DBS and bilateral pallidotomy in terms of movement disorder scale score in (a) with no significant overall publication bias (Egger’s test: p=0.31) and complication rates in (b) with no significant overall publication bias (Egger’s test: p=0.55).

A random effects model of total number of complications reported demonstrate an incidence of complications in DBS as 0.40 [0.19, 0.66] and incidence of complications after pallidotomy as 0.16 [0.03, 0.53] with a chi-square test for subgroup difference p-value of 0.14 and an I2 of 9.6% (Figure 3b). A funnel plot showed no publication bias with an Egger’s test of p=0.55 (Figure 4b).

A GRADE Summary of Findings table was generated to establish the fidelity of the reported complications and overall SD improvement, demonstrating low evidence for both complications and change in standardized motor score (Table 3).

**Table 3:**
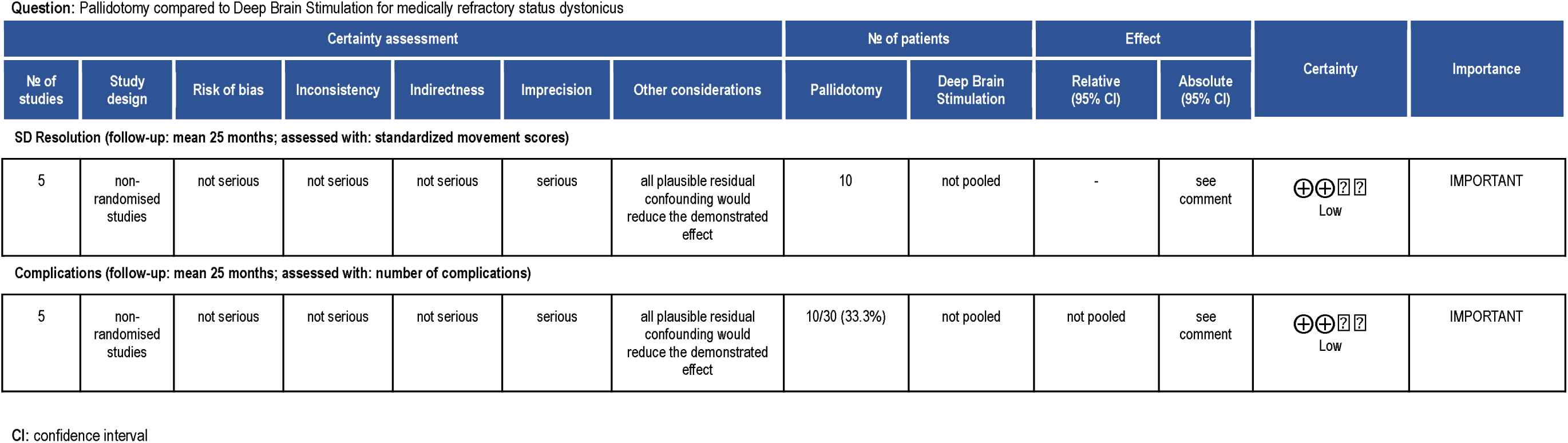
GRADE Summary of Findings Table.

Question: Pallidotomy compared to Deep Brain Stimulation for medically refractory status dystonicus
| Certainty assessment |  |  |  |  |  |  | No of patients |  | Effect |  | Certainty | Importance |
| --- | --- | --- | --- | --- | --- | --- | --- | --- | --- | --- | --- | --- |
| No of studies | Study design | Risk of bias | Inconsistency | Indirectness | Imprecision | Other considerations | Pallidotomy | Deep Brain Stimulation | Relative (95% CI) | Absolute (95% CI) |  |  |
| SD Resolution (follow-up: mean 25 months; assessed with: standardized movement scores) |  |  |  |  |  |  |  |  |  |  |  |  |
| 5 | non-randomised studies | not serious | not serious | not serious | serious | all plausible residual confounding would reduce the demonstrated effect | 10 | not pooled | - | see comment | ⊕⊕⊗⊗<br>Low | IMPORTANT |
| Complications (follow-up: mean 25 months; assessed with: number of complications) |  |  |  |  |  |  |  |  |  |  |  |  |
| 5 | non-randomised studies | not serious | not serious | not serious | serious | all plausible residual confounding would reduce the demonstrated effect | 10/30 (33.3%) | not pooled | not pooled | see comment | ⊕⊕⊗⊗<br>Low | IMPORTANT |

CI: confidence interval

## Discussion

This study aims to compare the efficacy of pallidotomy to DBS for medically refractory SD outcomes of standardized motor symptom score and complication rates. Given the rarity of the condition, there is a paucity of literature concerning the neurosurgical benefits and ramifications of these treatments.

There were no significant demographic differences except for the average age of dystonia onset, which was 9.02 ± 10.72 for DBS and 1.51 ± 1.24 for pallidotomy with a p<0.001 (Table 1). There is no evidence in the literature that age of dystonia onset influences SD treatment outcomes[4]. The results of the meta-analysis indicate that there is no statistically significant difference in standardized movement disorder score and complication rate between groups (Figure 3). It is important to note that every complication reported was included in the meta-analysis, which artificially elevated DBS complications with hardware-related side effects. The well-documented neurological side effects of pallidotomy were not reported in the included studies[37–39]. Neurological side effects were reported and found to be limited given the sample size and technical improvement in radiofrequency pallidotomy over the span of included studies.

Further subgroup analysis by etiology was not possible given the limited number of participants as well as the limited number of SD etiology within each surgical group (Table 1). A decision was made to include case reports in this overall analysis given the limited number of reported SD cases treated with neuromodulation. Literature review of the neurosurgical treatment of SD yielded over forty case reports with fewer than four subjects and reports, which would lower the quality of evidence if included within the meta-analysis[30]. An individual patient meta-analysis was not possible because the data collected involved the baseline movement disorder scale score while in SD and the latest follow-up measurement available after neurosurgical intervention.

If our methodology allowed, eleven further SD case reports would have been included, all of which were treated with DBS except for one pallidotomy and one combined DBS and pallidotomy case[35, 40–47]. The SD etiologies for DBS were as follows: four cerebral palsy (CP), three *PKAN* mutations, one primary dystonia (*DYT1*-negative), one *PANK2* mutation, one pyrexia due to an unknown cause, one primary segmental craniocervical dystonia, and one generalized dystonia. For pallidotomy alone, the patient had primary CP. All interventions led to significant improvement in overall clinical resolution of SD at short and medium length follow up visits with minimal complications. In one patient with Batten’s disease, however, DBS without failure or removal did not fully resolve SD and radiofrequency pallidotomy was utilized to resolve clinical SD. Overall, these case reports uphold the efficacy of DBS found in the included studies and demonstrate the usefulness of pallidotomy in the setting of suboptimal DBS.

This study reports four cases of SD episodes after DBS failure or hardware removal. Further studies are needed to be understand the risk of SD recurrence in these patient populations after DBS implantation. Discussions should be conducted by a multidisciplinary team based on the individual and their corresponding risk factors to minimize risk of these adverse events.

To the best of our knowledge, this is the first meta-analysis to directly compare DBS and pallidotomy for SD. There are systematic reviews and meta-analyses that demonstrate the efficacy of DBS for the treatment of medically refractory dystonia, but none address SD treatment nor the etiological subtypes with respect to surgical treatment of SD[6, 9].

This study has several limitations. Pooling case reports allows an appraisal of the existing literature but does not take surgeon and institutional preferences and abilities in account among other uncontrollable factors. The extent of the meta-analyses was curbed by the inclusion criteria and limited studies regarding the condition. The studies included were isolated case series except for one comparative cohort study, weakening the evidentiary strength. This is exemplified in the low level of evidence demonstrated in the summary of findings table for both complications and SD resolution (Table 3). While a standardized mean difference should correct for any differences in score used, a different version of a standardized motor score was used in three out of the five included studies, introducing more variability into the statistical comparisons. While there are only thirty-nine patients included, this study better contextualizes the techniques in global neurosurgery practice where patients may not have access to consistent neurological follow-up or longitudinal care involved in DBS programming, battery replacements and malfunction troubleshooting. This could make pallidotomy a viable alternative in such settings or those that have the same constraints applied.

## Conclusion

Based on the patients in our review, there was no difference in standardized motor score between pallidotomy and DBS for SD resolution, but the results are underpowered. There were differences in complication rates that did not reach clinical significance, as most for DBS involved small device complications. This suggests that pallidotomy may be a valid alternative for SD treatment in locations where DBS is unavailable or long-term neurological follow-up is limited.

## Data Availability

All data produced in the present study are available upon reasonable request to the authors

## Author Declaration

JK conceived of the project idea, performed the original investigation, search terms, performed initial and full-text screening along with data analysis and quality assessment and wrote the main manuscript text; DSR helped write the methods section and performed initial and full-text screening along with quality assessment; JAS performed the initial and full-texting screening and helped write the introduction section; MCK and RWB provided project oversight and both reviewed the original and final manuscript drafts as well as completed tables and figures.

